# Community-Based Helicobacter pylori Screening in High-Risk U.S. Populations: Protocol for a Mixed-Methods Study to Inform Gastric Cancer Prevention and Migration-Informed Risk Modeling

**DOI:** 10.1101/2025.08.28.25334491

**Authors:** Chul S. Hyun, Sarah Soyeon Oh, Sung Hwi Hong, Jae Il Shin

## Abstract

**Introduction:** Gastric cancer (GC) remains a persistent and underrecognized health disparity in the United States, disproportionately affecting immigrant communities from East Asia, Latin America, and other high incidence regions. Helicobacter pylori (H. pylori), a WHO Group 1 carcinogen and the leading modifiable risk factor for non cardia GC, remains largely unaddressed in U.S. screening guidelines, despite successful international eradication programs.

**Methods and Analysis:** This prospective, mixed methods study will enroll 2,000 adults aged 30 to 80 from Asian, Hispanic, and other high risk racial/ethnic backgrounds across the Greater New York area. Participants will be recruited through partnerships with trusted community organizations. Each will complete a multilingual survey and undergo H. pylori testing (urea breath test), with confidential follow up and referral. The survey will assess demographic, clinical, and KAP (knowledge, attitudes, practices) variables. A migration informed risk index will be developed using logistic regression and validated using k fold cross validation and bootstrap resampling.

**Ethics and Dissemination:** Approved by the Yale University IRB, with informed consent obtained in preferred languages. Results will be disseminated through community forums, peer reviewed publications, academic conferences, and policy briefings.

**Strengths and Limitations of This Study:**

- Culturally and linguistically sensitive screening and navigation strategies tailored to high-risk immigrant populations
- First U.S.-based effort to develop a migration-informed risk index for H. pylori using clinical, demographic, and KAP data
- Combines multilingual outreach and venue-based recruitment with predictive modeling to inform risk-stratified screening
- Enables detailed assessment of screening barriers and supports a multidimensional prevention framework
- Limitations include potential selection bias, limited geographic generalizability, and reliance on self-reported data; external validation will be necessary

## Introduction

Gastric cancer (GC) is a leading cause of cancer-related mortality worldwide and reflects persistent health disparities in the U.S., particularly among immigrants from East Asia, Latin America, and Eastern Europe. With approximately 1.1 million new cases and 770,000 deaths in 2020 alone, GC is projected to rise to 1.8 million cases and 1.3 million deaths by 2040.^1^ Although often viewed as a racial/ethnic disease, GC is fundamentally migration-linked, with risk concentrated in individuals from high-incidence regions. H. pylori infection, a major contributor to GC and typically acquired in childhood, is classified as a WHO Group 1 carcinogen.

Despite global eradication successes, the U.S. lacks national screening guidelines for GC or H. pylori, resulting in missed opportunities for prevention among immigrant communities.^2^ In contrast, countries like Japan and South Korea have implemented large-scale, government-supported screening programs tailored to population needs.^3^ Japan’s program increasingly incorporates endoscopic screening, which has demonstrated 2–8 times higher gastric cancer detection rates than radiography, and its reach continues to expand.^3^ South Korea’s National Cancer Screening Program (NCSP), launched in 2002, offers biennial screening to all adults over 40, allowing individuals to choose between endoscopy and X-ray.^3^ With participation rates exceeding 70% and a strong preference for endoscopy due to its greater sensitivity and low cost, South Korea’s model stands out as a scalable and effective approach to early gastric cancer detection.^3^

A recent multi-state analysis (CA, NY, TX, NJ, CT, GA, NM) revealed stark disparities in GC incidence and stage at diagnosis among Asian, Hispanic, and Black populations. Immigrant-dense areas shoulder a disproportionate burden, underscoring the need for community-based, migration-informed screening models that address both clinical risk and structural determinants.^4^ Increasingly, universal, interventional pilot studies such as Italy’s UNISCREEN pilot study, are illustrating the feasibility of capillary-based, population-level screening programs for detecting and/or preventing underdiagnosed conditions related to metabolic and cardiovascular risk.^5^ Such studies have demonstrated that point-of-care testing that is minimally invasive, and in alignment with community-embedded screening protocols integrating demographic, clinical, and behavioral data, may be an approach directly applicable to infection-based cancer prevention strategies like *H. pylori* screening among high-risk immigrant populations in the U.S.^5^

Similarly, recent efforts evaluating artificial intelligence (AI) in breast cancer screening through Australia’s national BreastScreen program demonstrated the feasibility of applying novel technologies in real-world, resource-limited settings. These models suggest that frameworks prioritizing clinical validity, early detection, and integration of population-based screening into existing public health infrastructures may be well-suited for *H. pylori* screening among high-risk immigrant populations in the United States.^6^ In rural Mexico, the SMART-SCREEN study has demonstrated that smartphone-based, population-level mental health screenings focused on suicide prevention can achieve significant community outreach. As a digital tool, it enables large-scale screening across geographically and socioeconomically diverse populations.^7^ This model illustrates that culturally tailored, community-based screening programs grounded in a migration-informed approach are not only feasible but also highly effective across a range of health domains.

Building on these perspectives, this study aims to address the lack of scalable, culturally tailored infection screening models for high-risk immigrant populations by piloting a community-based H. pylori screening program. Its innovation lies in the development and validation of a migration-informed risk index using clinical, demographic, and behavioral data. Inclusion of KAP variables offers a unique lens into behavioral drivers of infection and patterns of engagement with care.

## Methods and Analysis

### Study Objectives and Design

#### Primary Objective

To generate data for the development and internal validation of a migration-informed H. pylori risk index tailored to high-risk immigrant populations for targeted GC prevention.

#### Supporting Aims

- Implement a community-based H. pylori screening and treatment initiative
- Administer a multilingual survey capturing demographic, clinical, and KAP variables
- Establish referral pathways for H. pylori-positive individuals
- Evaluate screening uptake (≥30%), treatment completion (≥75%), and participant satisfaction
- Disseminate findings to researchers, policymakers, and community stakeholders

#### Study Design

This prospective, mixed-methods, community-based study will run from September 2025 to December 2026 and will enroll 2,000 adults aged 30–80 from high-risk Asian, Hispanic, and Black populations in the Greater New York area.

#### Sample Size Justification

The sample size of 2,000 was chosen to balance statistical power and feasibility. Assuming a conservative H. pylori prevalence of ∼30%, the study has >90% power to detect modest associations (OR 1.3–1.5) with key risk factors (α=0.05). Targeted recruitment will ensure meaningful representation from major subgroups—including Korean, Chinese, Japanese, Hispanic, and Black communities—with an expected sample size of 400–500 participants per major group. A small non-Hispanic White (NHW) reference group may be included for comparative purposes, where feasible. Projected recruitment targets by race/ethnicity are summarized in Table 1.

**Table 1.** Projected Recruitment Table.

| **Racial/Ethnic Group** | **Target Sample Size** | **Rationale** |
| --- | --- | --- |
| Korean American | 500 | High-risk group; strong community ties |
| Chinese American | 300 | High-risk group; large population in NY/NJ |
| Other Asian American | 200 | Includes Japanese, Vietnamese, Filipino, South Asian |
| Hispanic/Latino | 500 | High-risk; strong community partnerships |
| Black/African American | 300 | Emerging risk; underserved population |
| Non-Hispanic White (NH W) | 200 (optional) | Reference group for comparison |

### Study Population and Recruitment

#### Eligibility Criteria

- Age 30–80
- Belong to a moderate-to high-incidence stomach cancer ethnic group
- Able to understand English, Spanish, Korean, or Chinese
- Willing to provide informed consent

#### Recruitment Strategy

Participants will be recruited through community partners including Korean Community Services (KCS), ethnic associations, medical societies, and providers. Outreach will include multilingual flyers, cultural events, ethnic media, and lay health workers.

#### Data Collection Procedures

At health fairs and cultural venues, participants will:

- Provide informed consent
- Complete a 15-minute multilingual structured survey. The full structured survey instrument included as **Supplementary File 1**.
- Undergo urea breath testing for H. pylori

#### Survey Domains

1. Demographic and Migration History
2. Medical and Family History
3. Lifestyle and Risk Factors
4. Knowledge, Attitudes, and Practices (KAP)
5. Symptom Assessment
6. Post-Test Follow-Up and Referral

Participants testing positive for H. pylori will be notified confidentially within approximately three weeks and offered structured follow-up support. This includes printed bilingual resource guides, navigation assistance for accessing appropriate care, and follow-up calls to ensure linkage. Participants will also receive a standardized results letter to share with their primary care providers. The full linkage-to-care strategy by target community is outlined in Table 2.

**Table 2.** Post-Test Linkage-to-Care Strategy.

| **Community** | **Language** | **Navigator + Liaison** | **Resource Guide** | **Support at Result Disclosure** | **1–2 Month Follow-Up** |
| --- | --- | --- | --- | --- | --- |
| Korean | Korean/English | Yes | Provider list / FQHCs | Guide + explanation | Confirm care; assist if needed |
| Chinese/Asian | Mandarin/Asian | Yes | Same | Same | Same |
| Hispanic/Latino | Spanish/English | Yes | Same | Same | Same |
| Black/NH W | English | Yes | Same | Same | Same |

#### Primary Outcome

H. pylori prevalence

#### Secondary Outcomes

Screening uptake, treatment adherence, risk factor prevalence, participant satisfaction

#### Analytic Plan

- Logistic regression for risk factor analysis and index development
- Internal validation via k-fold cross-validation and bootstrapping
- AUC-ROC for discrimination; calibration plots for model fit
- Descriptive/inferential statistics (paired t-test, McNemar’s test)

#### Risk Index Development

Score weights will be derived from multivariable models including demographic, clinical, and KAP data.

### Ethics and Dissemination

#### Data Management and Ethics

##### Data Storage

All data will be stored on secure Yale servers. Surveys will be de-identified and coded; linking files stored separately. Paper files locked. HIPAA standards enforced.

##### Ethical Oversight

Approved by the Yale University IRB. Informed consent will be obtained. Deviations and problems reported per IRB protocol.

## Conclusion

This study will establish a scalable model for H. pylori screening in immigrant populations, surface structural and behavioral barriers to care, and inform future national screening policies through migration-informed risk tools and implementation strategies. Importantly, the risk index developed through this study may be externally validated and adapted for use in other U.S. cities with large immigrant populations, providing a foundation for broader implementation and comparative effectiveness research.

By directly responding to community needs, this study will pilot a culturally tailored *H. pylori* screening program grounded in migration-informed public health. It will integrate clinical, demographic, and behavioral data to develop and validate a practical risk index designed for real-world community settings. Aligned with broader efforts to advance risk-based strategies for early detection and prevention, the model is intended to be scalable to cities with large immigrant populations across the country; ultimately generating insights to inform clinical practice, guide public health policy, and promote equitable access to screening and early intervention. Findings will also support future NIH or PCORI-funded trials comparing clinic-vs. community-based screening and development of a clinically integrated risk index.

This study is particularly timely given the persistent and disproportionate burden of gastric cancer among first-generation immigrants from high-incidence regions. Unlike conditions such as hepatitis B or tuberculosis, where community-based models have led to measurable improvements in screening and care engagement, gastric cancer prevention efforts in the U.S. have remained largely hospital-centered.^2^ By embedding *H. pylori* screening within a community-driven, migration-informed framework, this study offers a practical model for aligning prevention strategies with population-specific risk while promoting equity in early detection and access to care.^2^

### Dissemination Plan

Results will be disseminated via:

- Peer-reviewed journals
- National conferences (ACG, DDW, AACR)
- Policy briefings (CDC, USPSTF)
- Community forums

### Timeline

- June 2025–Dec 2026: Recruitment/screening
- Jan–Mar 2027: Follow-up
- Apr–Jun 2027: Data analysis and dissemination

## Data Availability

No data used in study.

## Contributors

Concept and design: Jae Il Shin, Chul S. Hyun, Sung Hwi Hong

Acquisition, analysis, or interpretation of data: Chul S. Hyun

Drafting of the manuscript: Chul S. Hyun, Sarah Soyeon Oh

Critical revision of the manuscript for important intellectual content: Jae Il Shin, Chul S. Hyun, Sung Hwi Hong

Statistical analysis: Chul S. Hyun, Sung Hwi Hong

Administrative, technical, and material support: Jae Il Shin, Chul S. Hyun, Sung Hwi Hong

Supervision: Jae Il Shin, Chul S. Hyun

