## Supplemental File for "Community-Based Helicobacter pylori Screening in High-Risk U.S. Populations: Protocol for a Mixed-Methods Study to Inform Gastric Cancer Prevention and Migration-Informed Risk Modeling"

**Helicobacter pylori Screening and Gastric Cancer Risk Survey
 Participant Questionnaire**

**Section 1: Demographic Information**

**1. What is your age?**

**- Answer: _______ years**

**2. What is your sex?**

**- ☐ Male**

**- ☐ Female**

**3. Where were you born? (Country of birth)**

**- Answer: __________________**

**4. If born outside the U.S., how long have you lived in the U.S.?
- Answer: _______ years**

**5. What is your primary language spoken at home?
- Answer: __________________**

**6. How well do you speak English?
☐ Very well ☐ Well ☐ Not well ☐ Not at all**

**7. What is your highest level of education completed?
☐ Less than high school ☐ High school diploma ☐ Some college ☐ College graduate ☐ Graduate degree**

**8. Do you have health insurance?
☐ Yes ☐ No ☐ Unsure**

**9. What is your race?
- ☐ American Indian or Alaska Native**

**- ☐ Asian/Pacific islander**

**- ☐ Black or African American**

**- ☐ Hispanic, Latino, or Spanish origin**

**- ☐ Middle Eastern or North African**

**- ☐ White**

**- ☐ Other (please specify): ___________**

**Section 2: Medical and Family History**

**10. Have you ever been diagnosed with Helicobacter pylori infection?
- ☐ Yes
- ☐ No 🡪 Please move to Question 14 to continue
- ☐ Unsure**

**11. If your answer to Question 10 was yes, when were you diagnosed?**

**(Please write the year or approximate timeframe below.)
- Answer: ________________**

**12. If your answer to Question 10 was yes, how was your Helicobacter pylori infection diagnosed? (Check all that apply)**

**- ☐ Urea breath test
- ☐ Stool antigen test**

**- ☐ Blood test (serology)**

**- ☐ Endoscopy with biopsy**

**- ☐ Other (please specify): ___________
- ☐ I don’t know**

**13. If your answer to Question 10 was yes, have you ever received treatment for Helicobacter pylori infection?**

**- ☐ Yes, successfully treated**

**- ☐ Yes, but Helicobacter pylori was not eradicated after treatment by re-testing
- ☐ Yes, but I was not retested afterward
- ☐ No, I did not receive treatment
- ☐ Unsure**

**14. Do you have a family history of gastric cancer (stomach cancer)?
- ☐ Yes**

**- ☐ No 🡪 Please move to Question 17 to continue**

**- ☐ Unsure**

**15. If yes, who in your family had gastric cancer? (Check all that apply)**

**- ☐ Parent**

**- ☐ Sibling**

**- ☐ Grandparent**

**- ☐ Other (please specify): ___________**

**16. At what age was your family member diagnosed with gastric cancer?**

**- Answer: _______ years old**

**Section 3: Lifestyle and Risk Factors**

**17. Do you currently smoke?**

**- ☐ Yes
- ☐ No
- ☐ Former smoker**

**18. Do you drink alcohol?**

**- ☐ Never**

**- ☐ Occasionally (1–2 drinks per week)**

**- ☐ Frequently (3 or more drinks per week)**

**19. How often do you eat salty, pickled, or processed foods?**

**(e.g., pickled vegetables, salted fish, cured meats, etc.)
- ☐ Rarely/Never**

**- ☐ A few times per month**

**- ☐ A few times per week**

**- ☐ Daily**

**20. Have you ever been diagnosed with a stomach ulcer or gastritis?
- ☐ Yes**

**- ☐ No**

**- ☐ Unsure**

**21. Have you ever had an upper endoscopy (EGD) performed?
- ☐ Yes**

**- ☐ No 🡪 Please move to Question 24 to continue**

**22. If your answer to Question 21 was yes, when was the upper endoscopy performed?**

**(Please write the year or approximate timeframe below.)**

**Answer: ______________________________**

**23. If your answer to Question 21was yes, why did you had the procedure?**

**(e.g., routine screening, due to symptoms, follow-up evaluation)**

**Answer: ______________________________**

**Section 4: Awareness & Knowledge about Helicobacter pylori**

**24. Prior to your screening, how familiar were you with Helicobacter pylori?**

**- ☐ Not familiar**

**- ☐ Somewhat familiar**

**- ☐ Very familiar**

**25. Where did you first hear about Helicobacter pylori? (Check all that apply)
- ☐ Physician**

**- ☐ Media (TV, internet, newspapers)**

**- ☐ Family or friends**

**- ☐ Other (please specify): ___________**

**26. Did you know Helicobacter Pylori can cause stomach cancer?**

**- Answer: ________________________________**

**27. Did you know Asians and Hispanics had a greater chance of having stomach cancer than other races/ethnicities? - Answer: _____________**

**Section 5: Symptom Assessment**

**For each symptom below, please check the severity you have experienced in the past month on a scale from 0 (None) to 4 (Severe):**

**28. Abdominal pain or discomfort**

| **0** | **1** | **2** | **3** | **4** |
| --- | --- | --- | --- | --- |

**29. Bloating**

| **0** | **1** | **2** | **3** | **4** |
| --- | --- | --- | --- | --- |

**30. Nausea**

| **0** | **1** | **2** | **3** | **4** |
| --- | --- | --- | --- | --- |

**31. Loss of appetite**

| **0** | **1** | **2** | **3** | **4** |
| --- | --- | --- | --- | --- |

**32. Indigestion**

| **0** | **1** | **2** | **3** | **4** |
| --- | --- | --- | --- | --- |

**33. Frequent burping**

| **0** | **1** | **2** | **3** | **4** |
| --- | --- | --- | --- | --- |
